# A quantitative analysis of Alzheimer’s Disease and construction of an early Alzheimer’s detection deep learning system (EADDLS) using MRI data via machine learning along with ADmod: spatiotemporal-aware brain-amyloidβ growth model, using deep encoder-decoder networks about MRI

**DOI:** 10.1101/2024.08.02.24311435

**Authors:** Naitik B Mohanty, Morteza Sarmadi

**Affiliations:** Massachusetts Institute of Technology, Department of Mechanical Engineering

**Keywords:** Neural Network, Medical Resonance Imaging, Machine Learning, Alzheimer’s Disease

## Abstract

Alzheimer’s disease (AD) presents a significant societal challenge, with no current cure and an increasing prevalence among older adults. This study addresses the pressing need for early detection by harnessing the potential of machine learning applied to longitudinal MRI data. The dataset, sourced from the Open Access Series of Imaging Studies (OASIS) project, comprises MRI records of 150 subjects aged 60 to 96, each scanned at least once. Notably, 72 subjects were classified as ‘Nondemented,’ 64 as ‘Demented,’ and 14 underwent a transition from ‘Nondemented’ to ‘Demented,’ forming the ‘Converted’ category. What we propose is to develop a machine learning sound model capable of predicting the progression of mild cognitive impairment, leveraging key biomarkers extracted from MRI data. The chosen biomarkers include years of education (EDUC), socioeconomic status (SES), Mini-Mental State Examination (MMSE), Clinical Dementia Rating (CDR), Estimated Total Intracranial Volume (eTIV), Normalized Whole Brain Volume (nWBV), and Atlas Scaling Factor (ASF). Prior work in the field is referenced, highlighting studies that predominantly focused on raw MRI data analysis. In contrast, this study introduces a unique approach by utilizing a curated set of biomarkers, allowing for a more targeted and potentially interpretable model. Machine learning models such as Logistic Regression, Support Vector Machine, Decision Tree, Random Forest Classifier, and AdaBoost are employed, with performance measured using established metrics. Information about severity and state are stored during the EADDLS module and used for ADmod. ADmod uses the stored MRI data during the EADDLS module to model the growth of amyloid β build-up in the brain using convolution, resulting in both generalizable approaches and patient-specific approaches. There have been numerous mathematical instantiations to model amyloid β build-up using partial differential equations (or PDEs), these however have remained unincorporated due to prolonged runtimes and storage limitations along with those of pre-set conditions. We propose a novel amyloid β growth model using deep encoder-decoder networks in conjunction with convolution. The study contributes to the growing body of research in early Alzheimer’s detection, offering insights, results, and a discussion of limitations. The conclusion outlines a unique approach, emphasizes the practical implementation of the proposed model, acknowledges limitations, and suggests avenues for further research. Early detection of AD can significantly better the patient’s quality of care and lead to future preventative or risk assessment measures.

## 1.1 Introduction

Alzheimer’s disease (AD) stands as a formidable societal challenge, marked by its absence of a cure and a rising prevalence among older adults. The urgency for early detection has never been more pronounced, prompting researchers to explore innovative approaches to harness the potential of machine learning. This study responds to the critical need for early identification by delving into longitudinal MRI data, aiming to develop a robust machine learning model capable of predicting the progression of mild cognitive impairment. The dataset under scrutiny is derived from the Open Access Series of Imaging Studies (OASIS) project, encompassing MRI records of 150 subjects aged between 60 to 96, each subjected to at least one scan. The subjects are classified into ‘Nondemented,’ ‘Demented,’ and a distinctive ‘Converted’ category, comprising 14 individuals who transitioned from ‘Nondemented’ to ‘Demented.’ This unique classification sets the stage for a nuanced exploration of the disease’s progression.

This study proposes a machine-learning model that incorporates key biomarkers extracted from MRI data. These biomarkers encompass a spectrum of factors including years of education (EDUC), socioeconomic status (SES), Mini-Mental State Examination (MMSE), Clinical Dementia Rating (CDR), Estimated Total Intracranial Volume (eTIV), Normalized Whole Brain Volume (nWBV), and Atlas Scaling Factor (ASF). The deliberate selection of these biomarkers represents a departure from conventional approaches that predominantly focused on raw MRI data analysis. This curated set of biomarkers aims to provide a more targeted and potentially interpretable model for predicting the progression of mild cognitive impairment in individuals. The literature review within this introduction places the study in context by referencing prior work in the field. Notably, existing studies are highlighted for their emphasis on raw MRI data analysis. In contrast, this study takes a novel route by introducing a curated set of biomarkers, distinguishing itself through a more focused and potentially interpretable modeling approach. This departure from the conventional path underscores the study’s commitment to pushing the boundaries of current research in Alzheimer’s detection.

A variety of machine learning models are employed in this study, including Logistic Regression, Support Vector Machine, Decision Tree, Random Forest Classifier, and AdaBoost. The performance of these models is rigorously evaluated using established metrics, ensuring a comprehensive assessment of their efficacy in predicting the progression of mild cognitive impairment. This study’s contribution to the growing body of research in early Alzheimer’s detection is anticipated to be significant. The subsequent sections of the paper will delve into the insights gained, results obtained, and a nuanced discussion of the limitations encountered. Conversely, we purpose a novel spatio-temporal-aware brain amyloid β growth model using deep encoder-decoder networks in conjunction with a convolutional image to image regression architecture

## 2 Materials and Methods

### 2.1.1 Exploratory Data Analysis

In this section, we have focused on exploring the relationship between each feature of MRI tests and the dementia of the patient. The reason we conducted this Exploratory Data Analysis process is to state the relationship of data explicitly through a graph so that we could assume the correlations before data extraction or data analysis. Assisting us to understand the nature of the data and to select the appropriate analysis method for the model later.

The table is part of a dataset focused on Alzheimer’s disease detection using MRI data, where each row represents a unique subject identified by a Subject ID. The ‘Group’ column categorizes subjects as ‘Nondemented’ or ‘Demented.’ Information includes visit details, time delay for MRI scans (‘MR Delay’), demographic data (gender, handedness, age), education (‘EDUC’), and socioeconomic status (‘SES,’ some with missing values). Clinical assessments (‘MMSE’ and ‘CDR’) offer insights into cognitive function. MRI-related measurements encompass ‘eTIV,’ ‘nWBV,’ and ‘ASF.’ The ‘Group’ column is a potential target for predicting Alzheimer’s status based on these features.

According to a study at the Johns Hopkins School of Public Health; the link between cognitive decline and an elevated risk of Alzheimer’s disease (AD) dementia is often attributed to sleep apnea and inadequate sleep quality. Considering the higher prevalence of sleep apnea in men, the impact of this risk factor may be more pronounced in the overall context of men’s health (Mielke).

Normalized Whole Brain Volume is also found to be lower in the non-demented group rather than the demented group. This is shown in Figure 3.

**Figure 1:**
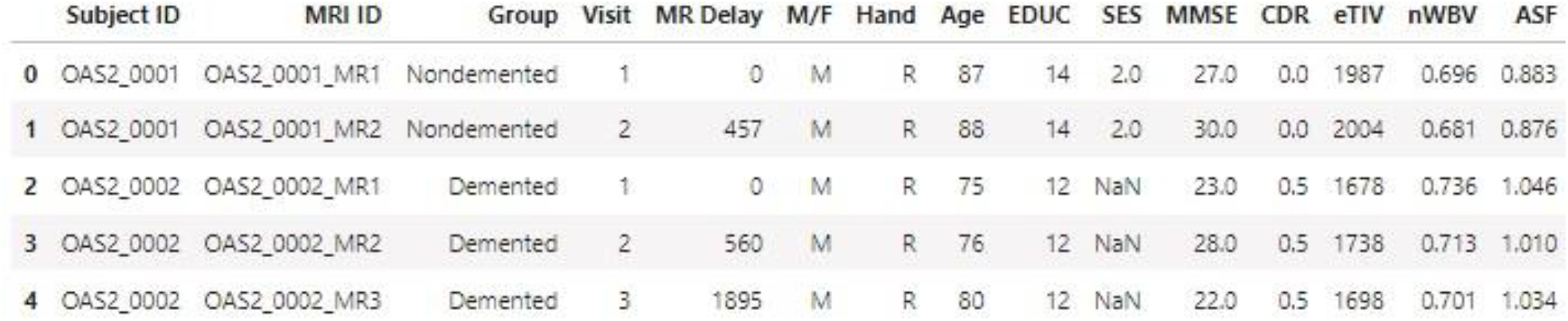
Table showing a subset of patient data storage.

**Figure 2:**
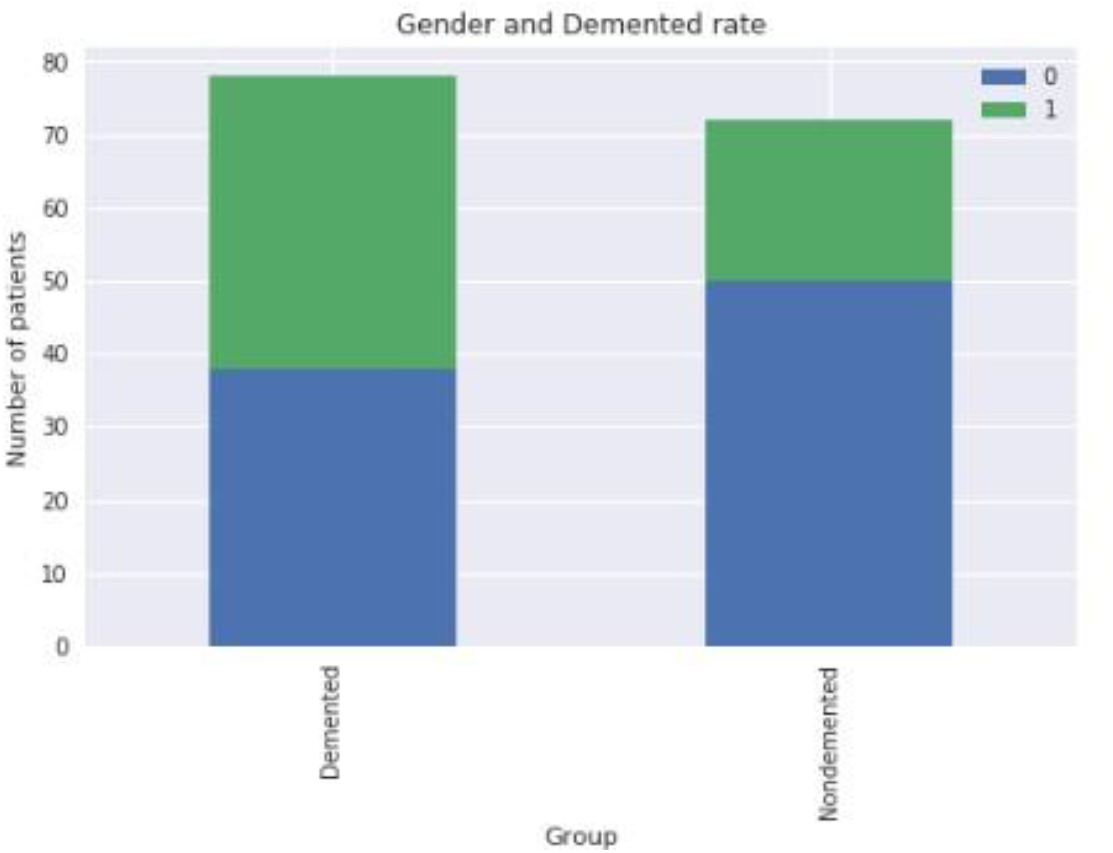
Demented vs Non-demented AD rate compared between males (0) and females (1). depicts that males are more likely to get AD than women, when demented, according to this dataset.

**Figure 3:**
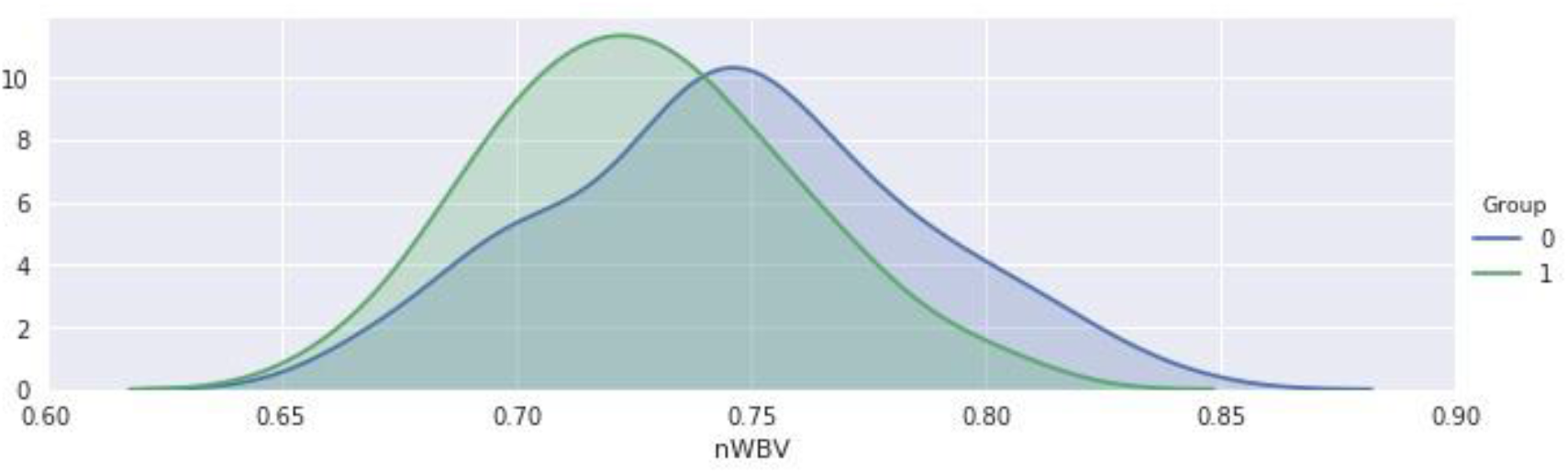
Non demented (1) nWBV vs Demented(0) nWBV. Graph indicating MRI Data, for which group (1) has overall a higher score of nWBV when compared to those of demented individuals.

**Figure 5:**
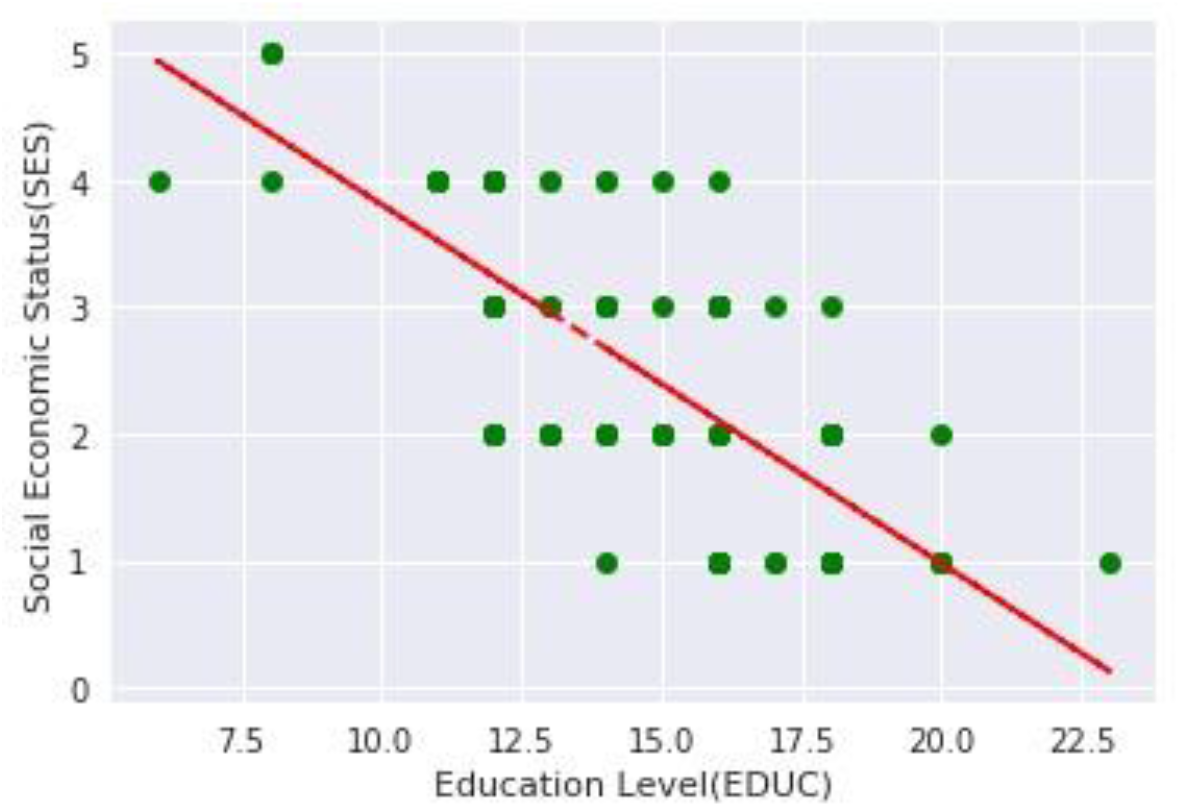
*EDUC v*.*s. SES*. Graph indicating a correlation between EDUC and SES.

**Figure 6:**
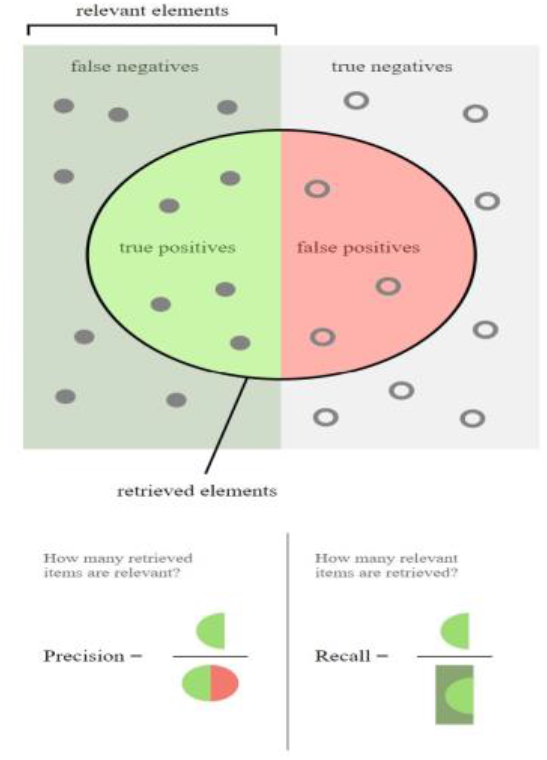
Visualization for model validation.

In summary of the data exploratory analysis; men have a higher likelihood of developing dementia, including Alzheimer’s Disease, compared to women. Demented patients generally exhibit lower levels of education in terms of years of schooling. Additionally, the demented group tends to have lower brain volume compared to the nondemented group, which is a common characteristic of neurodegenerative diseases. Moreover, there is a higher concentration of individuals aged 70-80 in the demented group, aligning with the typical age of onset for many forms of dementia, while the nondemented group includes a relatively lower proportion of individuals in this age range.

### 2.1.2 Data Pre-processing

In the given dataset, the SES (Social Economic Status) column has 8 missing values. To address this issue, two approaches are employed. In the first approach (5. A), the rows with missing SES values are simply dropped, resulting in a dataset without any missing values in the SES column. The new dataset, named df_dropna, consists of 72 instances in the “0” group and 70 instances in the “1” group. In the second approach, missing SES values are imputed by the corresponding median values based on the education level (EDUC), as SES is treated as a discrete variable. A scatter plot between EDUC and SES is drawn, and the imputation is performed manually using the median SES values for each education level.

This is shown in Figure 4.

The completeness of the data is confirmed, with all 150 data points utilized. Subsequently, the dataset is split into train/validation/test sets for both the imputed dataset and the dataset after dropping missing value rows. Feature scaling is applied to normalize the data.

A 5-fold cross-validation is conducted to determine the best parameters for different models, including Logistic Regression, SVM, Decision Tree, Random Forests, and AdaBoost. The tuning parameters are optimized based on accuracy. Finally, the performance metrics, including accuracy, recall, and AUC, are compared for each model. The entire process aims to evaluate and compare the impact of the chosen approaches (dropping vs. imputing missing values) on the performance of machine learning models using the given dataset.

### 2.1.3 Model Construction and Testing

We use the area under the receiver operating characteristic curve (AUC) as our main performance measure. We believe that in the case of medical diagnostics for non-life threatening terminal diseases like most neurodegenerative diseases, it is important to have a high true positive rate so that all patients with Alzheimer’s are identified as early as possible. But we also want to make sure that the false positive rate is as low as possible since we do not want to misdiagnose a healthy adult as demented and begin medical therapy. Hence AUC seemed like an ideal choice for a performance measure. We will also be looking at accuracy and recall for each model.

In the logistic regression model, the parameter C, representing the inverse of regularization strength, is tuned over the range [0.001, 0.1, 1, 10, 100]. The performance metrics, including accuracy, recall, and AUC, are evaluated using 5-fold cross-validation. The best-performing model is then selected based on the parameters that yield the highest accuracy on the validation set. The logistic regression model with imputation outperforms the one without imputation, demonstrating better accuracy (88.95% vs. 85%) and recall (85% vs. 80%). Moving on to the SVM model (6. C), the hyperparameters C, gamma, and kernel type are tuned over predefined ranges. The best-performing SVM model, determined through cross-validation, achieves an accuracy of 91.58% on the test set with a recall of 80%. The selected parameters include C=100, gamma=0.1, and the radial basis function (RBF) kernel.

### 2.1.3 Results of the Early Detection Module

In the decision tree model, the maximum depth is tuned over the range [1, 2, …, 8]. The best decision tree model is chosen based on cross-validated accuracy, resulting in a maximum depth of 1. The test set performance indicates an accuracy of 91.58% and a recall of 75%. The random forest classifier is optimized by tuning the number of trees (n_estimators), the maximum number of features considered at each split (max_features), and the maximum depth of the tree (max_depth). The best random forest model, with parameters M=14, d=5, and m=7, achieves a test set accuracy of 94.21% and a recall of 90%.

Finally, the AdaBoost model (6.F) is fine-tuned by adjusting the number of trees (M) and the learning rLRe (lr). The best AdaBoost model, with M=2 and lr=0.0001, exhibits a test set accuracy of 94.21% and a recall of 75%. Feature importance analysis reveals the contribution of each feature in the selected models. In summary, the models are systematically tuned and evaluated, with the random forest classifier demonstrating the highest accuracy and recall on the test set among the models considered. The performance comparison provides insights into the effectiveness of different machine learning algorithms in predicting the target variable based on the provided features. These results are represented in Figure 7.

**Figure 7:**
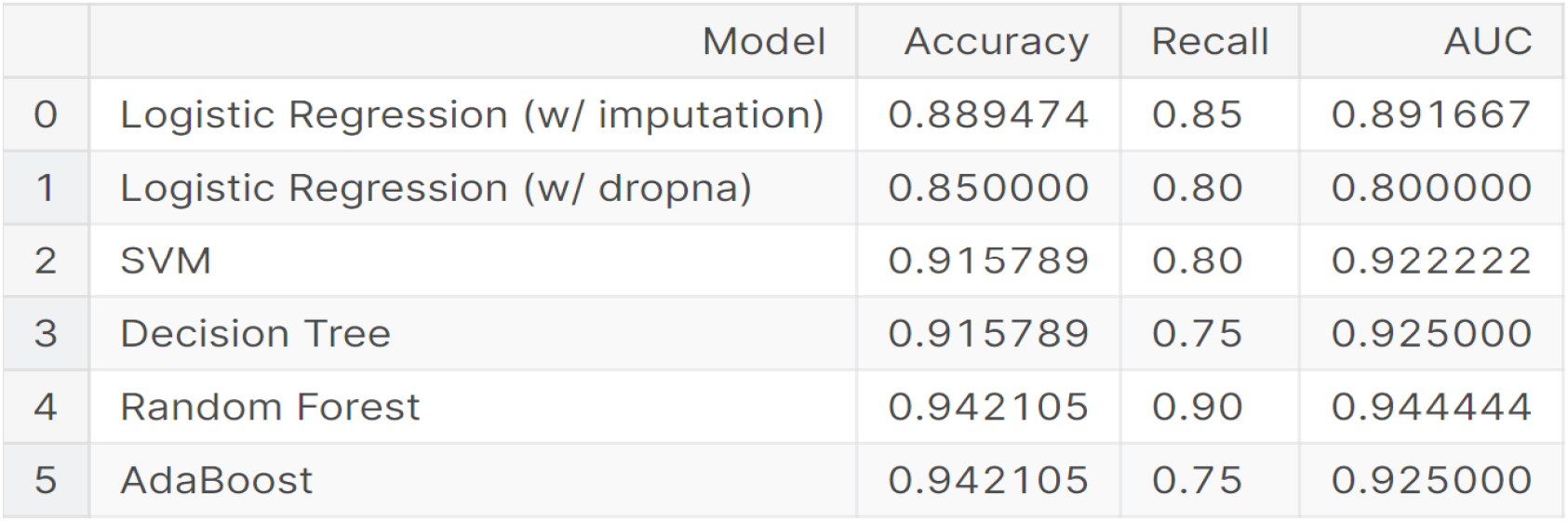
Results of testing Early Alzheimer’s Detection deep learning system.

This model also achieved a higher average accuracy than those previously tested. This is shown in Figure 8 below.

**Figure 8:**
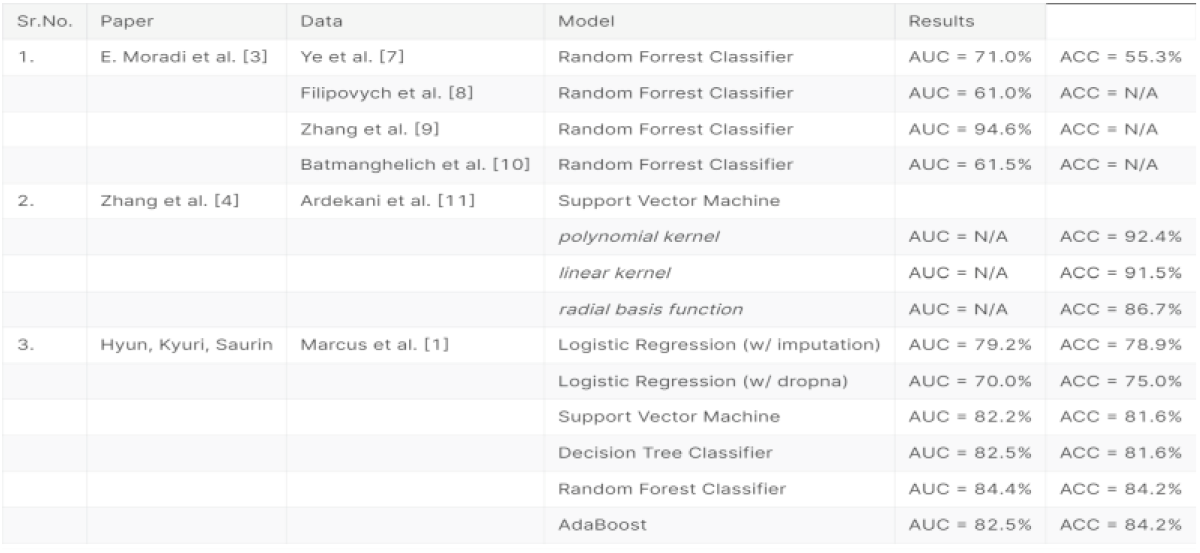
Previous early detection models. This table shows the accuracy of the previous study’s models.

### 2.2.1 Data Curation and Masking

We utilize a dataset from the OSASIS open source database, as published in the Journal of Nuclear Medicine. This dataset comprises 8 positron MRI scans of patients. MRI is instrumental in detecting amyloid β plaques in the brain.

### 2.2.2 Synthetic Plaque Generation

To augment the number of training samples for deep learning, synthetic plaque seed locations were randomly generated within the patient’s anatomy. This approach helps the neural model learn plaque growth patterns across various anatomical conditions. A total of one hundred synthetic plaques were created within the brain geometries of the eight patients. The randomized “plaque seeds” are produced within a NIfTI mask, which simplifies the scan by merging different segmentations. Seeds that are too close to the brain geometry are excluded to avoid complications in diffusivity outside the brain. This exclusion is based on an approximation of the central brain volume, derived from the average patient data obtained in section 2.1.1, “Exploratory Data Analysis.” An example of the NIfTI mask is shown in Figure 10 below.

**Figure 10:**
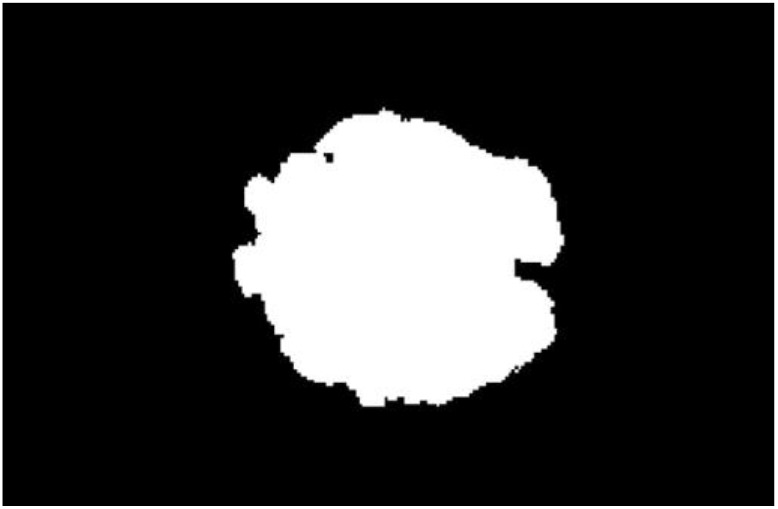
The central brain volume approximation utilized to restrict boundaries in which synthetic Aβ plaque seeds can be generated.

**Figure 11:**
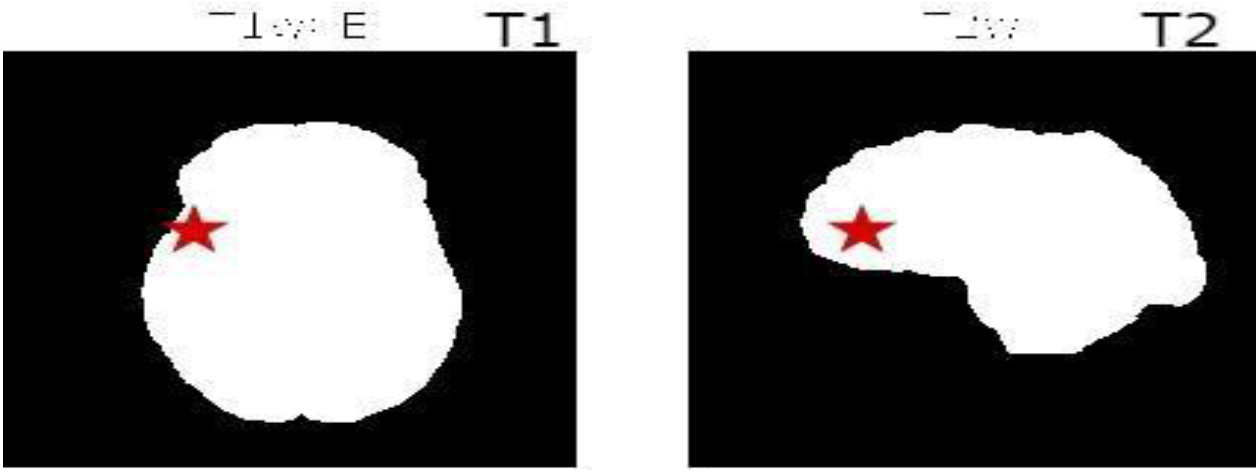
Montage of plaque seeds generated in XY-plane at varying Z values. Red asterisk represents the location of plaque seeds (x, y, z). These are two different patients T1 and T2. Where x ∈ [0, 240], y ∈ [0, 160], z ∈ [130, 170].

Additionally, examples of Aβ generated plaque seeds are shown below, derived from their corresponding MRI scans; identification of plaque location from MRI scans was done using an in-house CNN.

### 2.2.3 Deep Learning for Plaque Evolution

For convolution operations to effectively extract valuable features, the data must be arranged such that these features are spatially ordered. In ADmod, we use time-series images with temporally ordered predictions. However, exploiting the spatial properties of these images requires a different configuration than that used for tabular data in Section 2.2. While a hypothetical 1 × 3 convolution kernel could be applied to tabular data, it would sequentially process (x, y, z) and then (y, z, c), which is inadequate for spatial understanding. Therefore, tabular data must be transformed into a four-dimensional matrix suitable for convolution filters. These four dimensions are (i) height, (ii) channels, (iii) depth, and (iv) width. Ideally, interpolation to a 256 × 256 × 256 matrix is preferred; however, practical computational limits necessitate interpolation to a 64 × 64 × 64 matrix by normalizing each (x, y, z) point from 0 to 64 and assigning it the value of channel one. This results in a single-channel grayscale matrix.

Out of six patients and a total of 5,435 synthetic plaques, one patient is reserved for testing the model’s generalizability to unseen brain geometries. For the remaining five patients, with 100 plaques per patient and 10 initial starting times, 5,000 samples span days 0 through 500. From these, two four-dimensional matrices each containing 4,500 samples are created: 1) an input matrix with samples from day 0 to 450, and 2) an output matrix with samples from day 10 to 500. The encoder-decoder model’s fixed depth requires the training data to have a consistent interval.

### 2.2.4 Parameter Organization and Model Architecture

To predict future plaque conditions, it is crucial to understand both the biophysical characteristics and the patient’s brain geometry. Deep learning is employed to enhance the integration of these traits and to comprehend the underlying spatiotemporal features, providing an advantage over traditional PDE-based methods. ADmod uses an innovative encoder-decoder neural network for image-to-image regression, enabling the prediction of future plaque states. The model architecture consists of a 33-layer deep encoder-decoder network with 36,500 learnable parameters. The architecture features 3D Convolution → Batch Normalization → Swish → 3D Max Pool operations, followed by Transposed Convolution → Batch Normalization → Swish operations.

Hyperparameters were iteratively tuned based on training dataset performance. This iterative optimization approach prevents overfitting while maintaining high performance. The learning rate followed a piecewise schedule, being multiplied by a decay factor each epoch. This initial learning rate offered a balanced tradeoff between convergence and precise training. The decay was implemented to enhance network generalization and improve the learning of complex patterns during later training stages. The hyperparameters are detailed in Figure 12.

**Figure 12:**
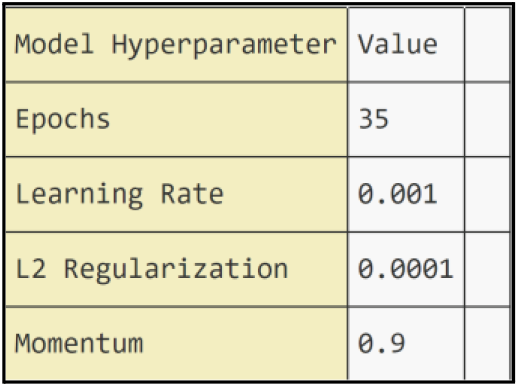
Table showing values of hyperparameters used. The type of hyperparameter is listed to the left and its specific value is listed in the left hand column.

### 2.2.4 Parameter Organization and Model Architecture

Key architectural modifications include replacing Rectified Linear Unit (ReLU) activation with Swish activation, incorporating batch normalization, and employing larger kernel sizes in initial convolutions. Swish activation has demonstrated superior performance over ReLU, albeit with higher computational demands. Batch normalization is integrated within the encoder-decoder framework between convolution and nonlinearity to expedite model convergence and reduce training duration. Larger kernel sizes, such as the 7 × 7 × 7 filter in the first convolution followed by smaller 3 × 3 × 3 filters later in the model, are adopted to capture a broader context of both the plaque surroundings and its edges.

### 2.2.5 Results of Growth Model Module

For evaluating ADmod, an image-to-image regression model, its performance is assessed using four common regression error metrics: root mean squared error (RMSE), relative root mean squared error (RRMSE), and relative squared error (RSE). These metrics quantify the disparity between predicted pixel intensities and ground-truth values across 900 image pairs representing plaque growth simulation. The input images cover days 0 to 450, while the output spans days 50 to 500. ADmod’s predictions are compared to mathematically predicted outputs. The evaluation on the test subset shows ADmod achieves an RMSE of 0.0204 ± 0.0001, RRMSE of 0.0013 ± 0.00001, and RSE of 0.3735 ± 0.0049.

Furthermore, the error assessment involves quantifying the agreement between ADmod-predicted and ground-truth simulations on the testing dataset. Each simulation is condensed into a single value by summing the pixel values in the image. These values are then paired, and linear regression is performed on the resulting 900 pairs. Dayeh et al.’s analysis is utilized to unveil systematic errors in the model by measuring agreement between ADmod and ground-truth predictions.

### 2.2.6 Visual and Quantitative Analysis

While the primary focus of this study is predicting plaque concentration, accurately reconstructing the surrounding patient geometry is also crucial for assessing the model’s success. This aspect is evaluated in the error analysis outlined in Section 2.2.5 and qualitatively demonstrated in Figure 15. The figure illustrates the reconstruction of patient geometry in a forward timestep, even amidst plaque growth, highlighting the model’s ability to capture the broader anatomical context.

**Figure 15:**
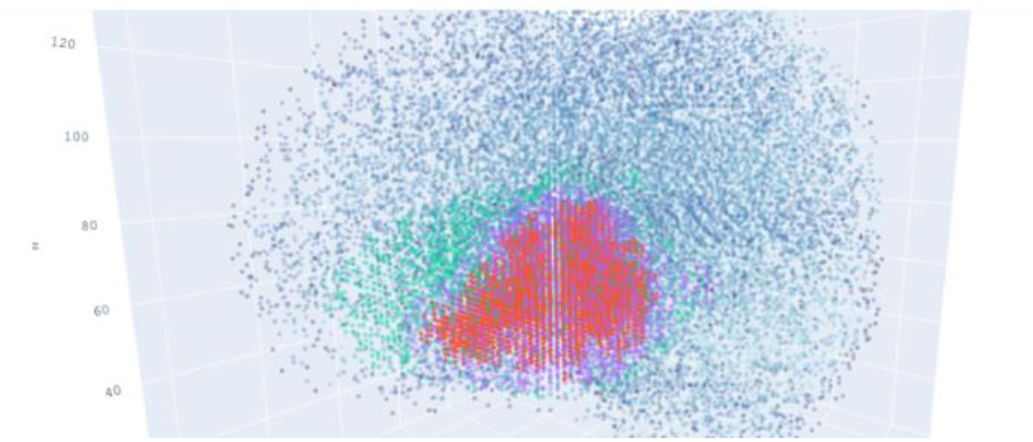
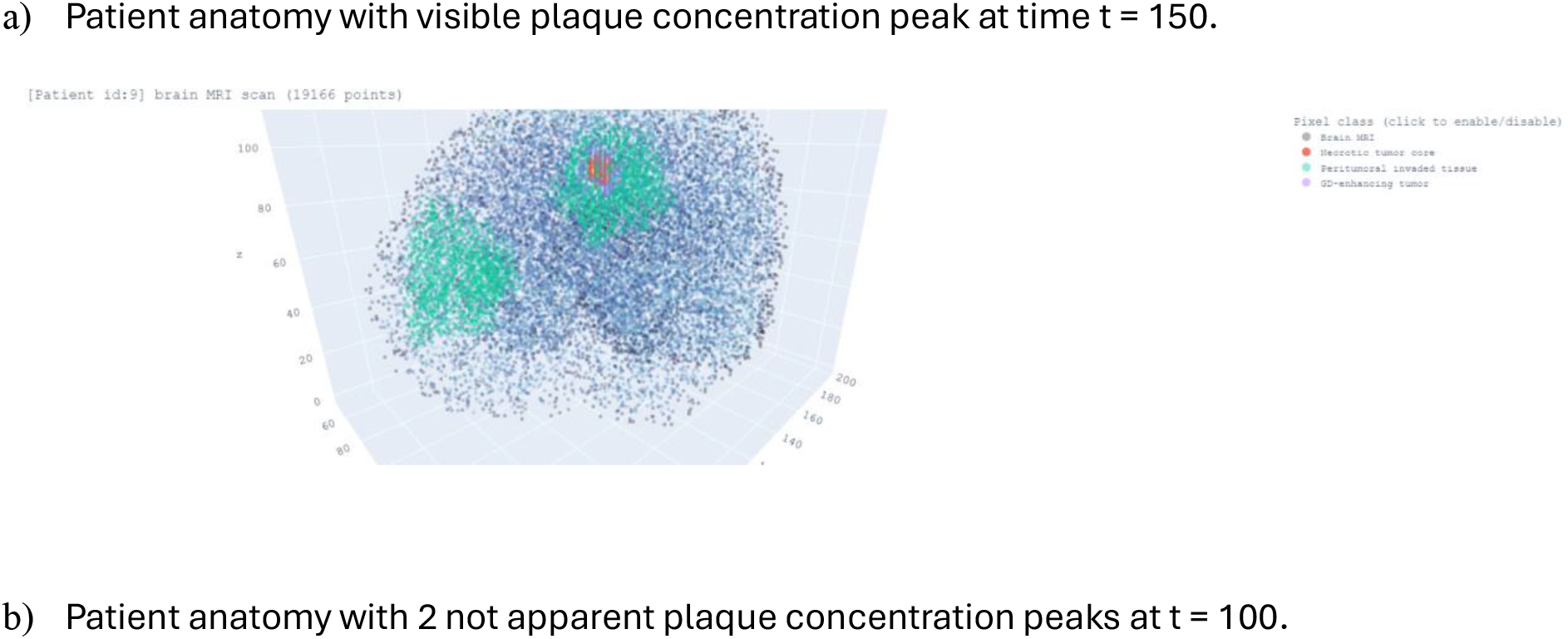
ADmod reconstructs surrounding patient geometry while growing plaque taking about 1.54 seconds per simulation.

## 3.0 Discussion

In summary, the unique aspect of our approach lies in incorporating key metrics such as the Mini-Mental State Examination (MMSE) and educational background into our model, thereby enhancing its ability to differentiate between normal healthy adults and those with Alzheimer’s disease. The MMSE, a gold standard in dementia assessment, adds a crucial dimension to our predictive model. This distinctive feature also lends flexibility to our approach, allowing it to be extended to other neurodegenerative diseases diagnosed through a combination of MRI features and cognitive tests. The Early Alzheimer’s Detection Deep Learning System can achieve up to 94% accuracy in detecting Alzheimer’s disease.

In terms of implementation, our primary focus was to bridge the gap between machine learning advancements and clinical applications. To facilitate this, we developed a web program using our algorithm, designed to be user-friendly for individuals regardless of their programming experience. Leveraging the Common Gateway Interface (CGI), our web application allows clinicians to input MRI results, biographic data, and other relevant patient parameters. The model, integrated into the web platform, then assists clinicians in identifying dementia, making our approach both accessible and impactful in a clinical environment.

However, it is crucial to acknowledge the limitations of our study. The size of the dataset poses challenges for implementing a more complex model, and further refinement in the data cleaning and analysis process could potentially lead to improved prediction rates. Additionally, variations in dataset characteristics might impact the generalizability of our model. This issue is evident in the perfect recall score of 1.0 for the SVM, highlighting the need for caution when applying the model to different datasets.

For future research, we emphasize the exploration of key factors contributing to dementia through sophisticated exploratory data analysis (EDA) with a larger sample size. This includes not only refining existing variables but also exploring novel dimensions such as generational grouping, brain tissue volume grading, and exam scores. Integrating insights gained from this process into data cleaning could enhance the model’s decision-making ability, potentially elevating the accuracy of our predictive model for early Alzheimer’s detection.

## Data Availability

All data produced in the present study are available upon reasonable request to the authors
All data produced in the present work are contained in the manuscript
All data produced are available online at Kaggle.com

https://sites.wustl.edu/oasisbrains/home/oasis-2/

## References

1. Marcus DS, Fotenos AF, Csernansky JG, Morris JC, Buckner RL. Open Access Series of Imaging Studies (OASIS): Longitudinal MRI Data in Nondemented and Demented Older Adults. Journal of cognitive neuroscience. 2010;22(12):2677–2684. doi:10.1162/jocn.2009.21407.

2. Marcus, DS, Wang, TH, Parker, J, Csernansky, JG, Morris, JC, Buckner, RL. Open Access Series of Imaging Studies (OASIS): Cross-Sectional MRI Data in Young, Middle Aged, Nondemented, and Demented Older Adults. Journal of Cognitive Neuroscience, 19, 1498–1507. doi:10.1162/jocn.2007.19.9.1498

3. Elaheh Moradi, Antonietta Pepe, Christian Gaser, Heikki Huttunen, Jussi Tohka, Machine learning framework for early MRI-based Alzheimer’s conversion prediction in MCI subjects, In NeuroImage, Volume 104, 2015, Pages 398–412, ISSN 1053-8119, 10.1016/j.neuroimage.2014.10.002.

4. Zhang Y, Dong Z, Phillips P, et al. Detection of subjects and brain regions related to Alzheimer’s disease using 3D MRI scans based on eigen brain and machine learning. Frontiers in Computational Neuroscience. 2015;9:66. doi:10.3389/fncom.2015.00066.

5. Magnin, B., Mesrob, L., Kinkingnéhun, S. et al. Support vector machine-based classification of Alzheimer’s disease from whole-brain anatomical MRI. Neuroradiology (2009) 51: 73. 10.1007/s00234-008-0463-x

6. http://scikit-learn.org/stable/modules/preprocessing.html#imputation

7. Ye, D.H., Pohl, K.M., Davatzikos, C., 2011. Semi-supervised pattern classification: application to structural MRI of Alzheimer’s disease. Pattern Recognition in NeuroImaging(PRNI), 2011 International Workshop on. IEEE, pp. 1–4. 10.1109/PRNI.2011.12.

8. Filipovych, R., Davatzikos, C., 2011. Semi-supervised pattern classification of medical images: application to mild cognitive impairment (MCI). Neuroimage 55 (3), 1109–1119. 10.1016/j.neuroimage.2010.12.066

9. Zhang, D., Shen, D., 2012. Predicting future clinical changes ofMCI patients using longitudinal and multimodal biomarkers. PLoS One 7 (3), e33182. 10.1371/journal.pone.0033182

10. Batmanghelich, K.N., Ye, D.H., Pohl, K.M., Taskar, B., Davatzikos, C., 2011. Disease classification and prediction via semi-supervised dimensionality reduction. Biomedical Imaging: From Nano to Macro, 2011 IEEE International Symposium on. IEEE, pp. 1086–1090. 10.1109/ISBI.2011.5872590

11. Ardekani, B.A., Bachman, A.H., Figarsky, K., andSidtis, J.J. (2014). Corpus callosum shape changes in early Alzheimer’s disease: an MRI study using the OASISbraindatabase. BrainStruct.Funct. 219, 343–352. doi:10.1007/s00429-013-0503-0

12. Dayeh, Maher A et al. “A discrete mathematical model for the aggregation of β-Amyloid.” PloS one vol. 13, 5 e0196402. 23 May. 2018, doi:10.1371/journal.pone.0196402

13. Nafiseh Ghazanfari and Jeih-San Liow and Min-Jeong Kim and Raven Cureton and Adrian Lee and Carson Knoer and Madeline Jenkins and Jinsoo Hong and Jose A. Montero Santamaria and H. Umesha Shetty and Anthony Galassi and Paul Wighton and Martin Norgaard and Douglas N. Greve and Sami S. Zoghbi and Victor W Pike and Robert B Innis and Paolo Zanotti-Fregonara (2024). [11C]PS13 demonstrates pharmacologically selective and substantial binding to cyclooxygenase-1 (COX-1) in the human brain. OpenNeuro. [Dataset] doi: 10.18112/openneuro.ds004868.v1.0

